# Trajectories of posttraumatic stress and obsessive-compulsive symptoms over twelve months following Hurricane Helene

**DOI:** 10.64898/2026.05.18.26353502

**Authors:** Caitlin M. Pinciotti, Helen Pushkarskaya, Iasha Williams, Emily Olfson, Thomas G. Adams

**Author notes:** **Corresponding Author:** Caitlin M. Pinciotti, PhD, One Baylor Plaza, Houston, TX USA 77030.

## Abstract

Separate research has evaluated trajectories of posttraumatic stress symptoms (PTSS) and obsessive-compulsive symptoms (OCS), but no study has evaluated OCS trajectories following trauma exposure nor combined PTSS/OCS trajectories. The present study evaluated combined PTSS/OCS trajectories among 585 survivors of Hurricane Helene, spanning three waves of data collection over 12 months. A 3-class solution was supported, including resilient (i.e., consistently low PTSS and OCS), chronic (i.e., elevated PTSS and OCS with gradual reduction over time), and moderate-yet-diverging (i.e., moderate elevations in PTSS and OCS with gradually declining PTSS and persistent and increasing OCS over time) classes. This study shows both overlap and differentiation in symptom trajectories from earlier research, with the moderate-yet-diverging trajectory suggesting unique OCS pathways distinct from PTSS.

## Introduction

Following trauma exposure, posttraumatic stress symptoms (PTSS) tend to follow one of four trajectories in the 12-to-24 months following trauma exposure (resilience, recovery, chronic, delayed).^1^ Although minimal research suggests trajectories of high-but-remitting, moderate-but-escalating, and low-and-stable for obsessive-compulsive symptoms (OCS),^2^ no study has accounted for the impact of trauma exposure on OCS trajectories. Given the impact of trauma exposure on the emergence and chronicity of OCS,^3–6^ this study evaluated the combined trajectories of PTSS and OCS over 12 months following Hurricane Helene.

## Methods

This study was reviewed and approved by the Baylor College of Medicine Institutional Review Board (#H-56103). A total of 585 participants impacted by Hurricane Helene were recruited to participate in a longitudinal self-report study involving three waves of data collection.^7^ The first wave recruited participants within 4-to-12 weeks of Hurricane Helene landfall in the U.S. (October 28, 2024 – January 18, 2025). Wave 2 surveys were automated to be sent out exactly three months after each participant completed their baseline survey, remaining open for 8 weeks total (January 22, 2025 – May 4, 2025); and Wave 3 began approximately 12 months after Hurricane Helene made landfall in the U.S. (regardless of when Wave 1 or 2 was completed), remaining open for 9 weeks total (October 2, 2025 – December 1, 2025).

PTSS and OCS were measured at each time point based on the PTSD Checklist for DSM-5 (PCL-5)^8^ and Padua Inventory-Washington State University Revision (PI-WSUR).^9^

Latent class trajectory analysis was conducted using the multlcmm function from the lcmm package in R 4.4.1 (code is available in supplementary materials). Models jointly included standardized PTSS and OCS as parallel longitudinal outcomes. A one-class model was first estimated with fixed effects of time, a participant-level random intercept, and linear links for both outcomes. Models with two through six classes were subsequently estimated using class-specific effects of time, participant-level random intercepts, and linear outcome links. To reduce the risk of local maxima, each multi-class solution was estimated using grid search with 50 random starts and 30 iterations per start, initialized from the one-class model.

Before fitting latent class models, individual symptom trajectories were screened for extreme within-person change. PTSS and OCS scores were z-standardized, and participants were flagged as potential trajectory outliers if the within-person range exceeded 3 standard deviations on either measure. Flagged cases were then visually inspected using both raw-score and standardized trajectory plots.

Model evaluation was based on convergence, log-likelihood, Akaike Information Criterion (AIC), Bayesian Information Criterion (BIC), sample-size adjusted BIC (SABIC), entropy, minimum class size, and clinical interpretability of the resulting trajectories. Likelihood-ratio improvements between adjacent class solutions were calculated as twice the change in log-likelihood. Approximate p-values were derived using a chi-square reference distribution based on changes in estimated parameters; however, these values were interpreted cautiously because standard likelihood-ratio assumptions are frequently violated in mixture models.

Following initial model comparison, two candidate solutions were retained for further evaluation. For each candidate model, posterior class probabilities were extracted and participants were assigned to the class associated with the highest posterior probability.

Classification quality was evaluated using entropy, mean maximum posterior probability, and class-size distributions.

To further assess model stability, bootstrap refitting procedures were conducted for each candidate solution. Bootstrap samples were generated by resampling participants with replacement while preserving all longitudinal observations within participant. Each bootstrap sample was assigned new participant identifiers so that resampled trajectories were treated as distinct individuals during model estimation. For each bootstrap sample, the one-class model was refit and used to initialize the corresponding multi-class solution. Fifty bootstrap samples were generated per candidate model, with 20 random starts and 50 grid-search iterations per bootstrap refit. Stability was summarized using the proportion of converged solutions, mean and standard deviation of entropy, and mean and standard deviation of the smallest class percentage across converged bootstrap refits. Class-specific symptom slopes were also examined descriptively within bootstrap samples by estimating mean symptom change across time within assigned classes.

Finally, to evaluate potential biases related to dropout, missingness and attrition were examined descriptively for each candidate solution. Overall missingness in PTSS and OCS scores was summarized by wave. To evaluate class-specific attrition, each participant’s expected longitudinal wave structure was reconstructed, and next-wave missingness was coded for each observed nonterminal wave. Retention was summarized as the percentage of participants retained at each wave relative to baseline class size. Next-wave missingness was additionally summarized as a function of latent class membership and current symptom severity. To evaluate whether dropout varied as a function of symptom burden, PTSS and OCS scores were divided into within-class tertiles, and next-wave dropout rates were examined separately across PTSS and OCS severity strata. Associations between latent class membership and next-wave missingness were evaluated using chi-square tests and interpreted as descriptive sensitivity analyses rather than primary inferential tests.

## Results

Inspection of individual trajectories identified three potential outliers (See Figure S1), characterized by unusually large within-person changes across waves (more than 3 *SD* from the sample means). These cases were excluded from the subsequent analyses.

Model fit indices generally favored the 3-class solution, with the 4-class solution emerging as a secondary candidate for further evaluation (Figure 1). Relative to the 2-class model, the 3-class solution showed improved fit across major information criteria (AIC = 2227.19, BIC = 2272.81, SABIC = 2231.60) while maintaining high classification quality (entropy = 0.885) and clinically interpretable class sizes (smallest class = 8.5%). Although the 4-class solution produced a modest improvement in log-likelihood, this improvement was limited relative to the increase in model complexity, and information criteria worsened compared to the 3-class solution (AIC = 2228.60, BIC = 2284.75, SABIC = 2234.03). Entropy also declined (0.831), and the smallest class decreased to 4.45% of the sample. Models with five or more classes showed further deterioration in classification quality and class stability; entropy declined substantially in the 5-class and 6-class solutions, and minimum class sizes became small (<6% of the sample), suggesting potential overfitting.

**Figure 1.**
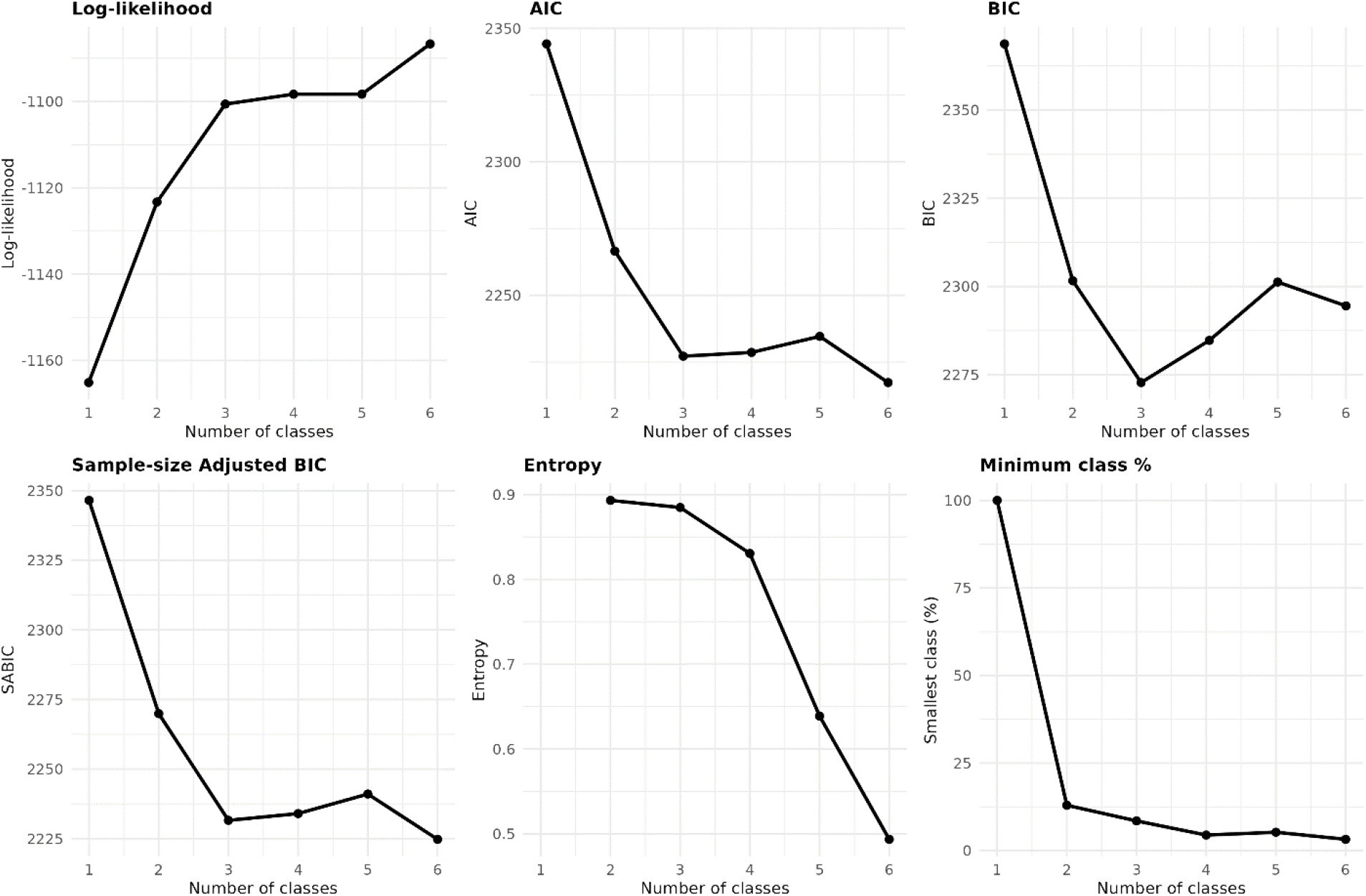
Model fit indices.

Bootstrap analyses supported the relative stability of the 3-class solution compared to the 4-class solution. The 3-class model demonstrated complete convergence across all bootstrap refits (50/50; 100%), consistently high entropy (mean = 0.90 ± 0.02), and relatively stable minimum class sizes (mean = 8.15% ± 2.10), suggesting robust class separation and limited sensitivity to sampling variation. In contrast, the 4-class solution showed lower convergence rates (47/50; 94%), substantially greater variability in entropy (mean = 0.81 ± 0.10), and more unstable minimum class sizes (mean = 6.59% ± 3.26), with 7 of 50 bootstrap samples yielding classes smaller than 3% of the sample and the smallest class reaching 1.6%. Taken together, these findings supported the 3-class solution as the more stable and parsimonious representation of longitudinal PTSS and OCS trajectories.

Descriptive analyses of class-specific symptom slopes across bootstrap refits were consistent with this pattern, with the 4-class solution showing greater variability in PTSS and OCS trajectory estimates than the 3-class solution, particularly for OCS slopes (SDs up to 0.09), suggesting reduced stability of class-specific longitudinal patterns in the 4-class model.

To evaluate the clinical interpretability of the candidate solutions, symptom trajectories were examined for the 3- and 4-class models (Figure 2A–B). Both solutions identified a resilient class (Class 2 in both models), representing the largest subgroup in the sample and characterized by consistently low PTSS and OCS following trauma exposure. Both solutions also identified a chronic class (Class 3 in the 3-class solution; Class 4 in the 4-class solution), characterized by elevated PTSS and OCS immediately following trauma exposure, with gradual symptom reduction over time. These two classes were broadly comparable across solutions.

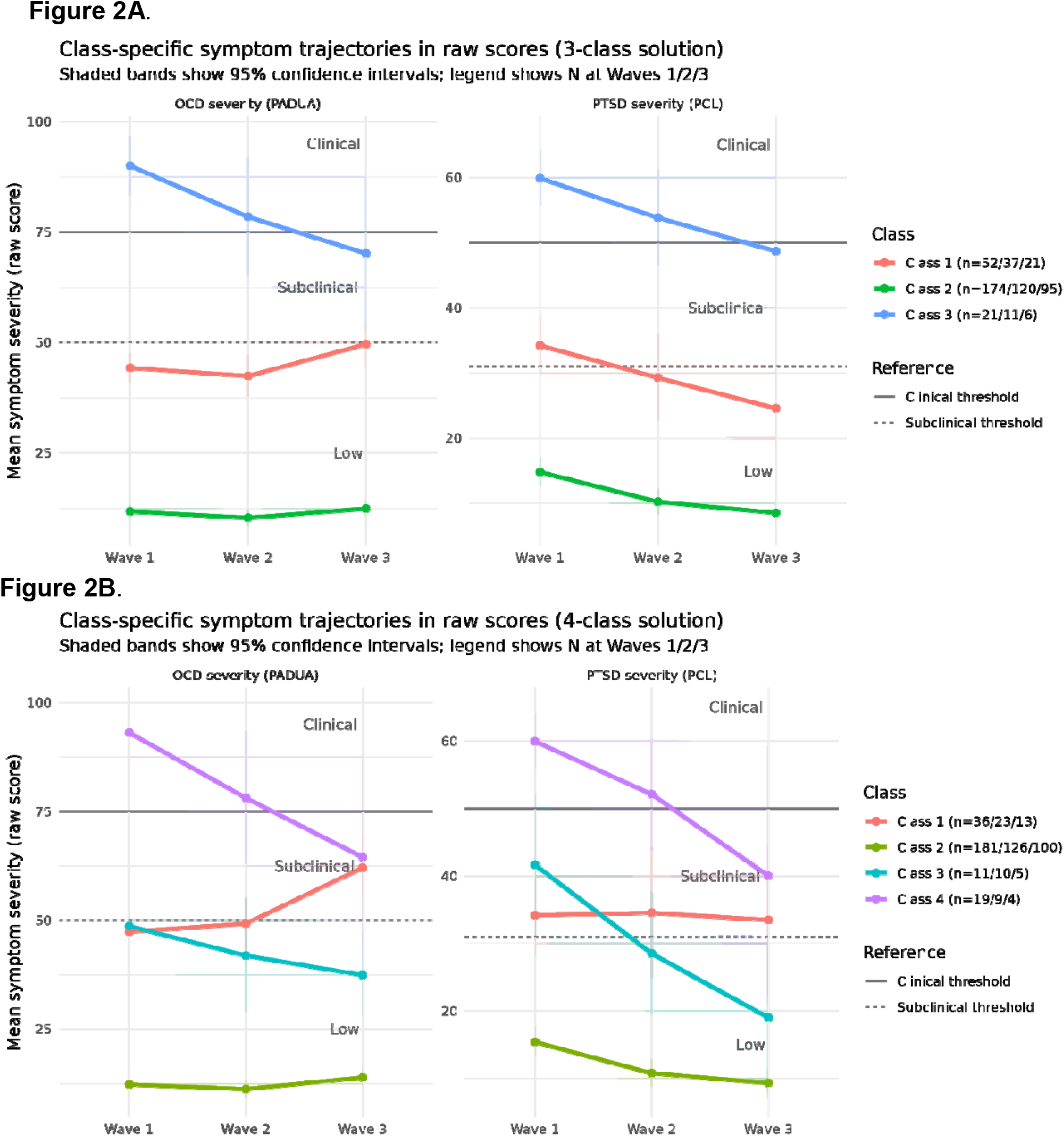

The 3-class solution additionally identified a moderate-yet-diverging class characterized by borderline subclinical elevations in both PTSS and OCS following trauma exposure. Within this class, PTSS gradually declined over time, whereas OCS persisted and showed a modest increasing trend. This pattern suggests differential longitudinal trajectories for PTSS and OCS following trauma exposure. The 4-class solution suggested potential heterogeneity within this intermediate subgroup, separating individuals with gradual improvement across both symptom domains from those with persistently elevated PTSS and increasing OCS over time. However, these additional classes were relatively small and less stable across bootstrap analyses, limiting confidence in the robustness of this finer-grained subdivision within the current sample.

Finally, missingness analyses indicated substantial attrition across waves, particularly in the symptomatic classes (Figure S2). In the 3-class solution, Wave 3 retention declined to approximately 40% in the moderate-yet-diverging class and 29% in the chronic class, compared to 55% in the resilient class. Similarly, in the 4-class solution, Wave 3 retention declined to approximately 36% in Class 1, 45% in Class 3, and 21% in the chronic Class 4, compared to 55% in the resilient class. This pattern raises the possibility that longitudinal trajectories may underestimate symptom persistence over time if individuals with more severe or chronic symptoms were less likely to remain in the study. However, within-class baseline symptom severity was generally comparable between participants who remained in the study and those who subsequently dropped out. For example, in the chronic class of the 3-class solution, baseline PTSS/OCS scores were similar among retained versus dropout participants (57.2/82.7 vs. 58.4/89.8). Likewise, in the moderate-yet-diverging class, retained and dropout participants showed comparable baseline PTSS (32.9 vs. 30.7) and OCS (42.8 vs. 44.8) levels. Similar patterns were observed across classes in the 4-class solution, although somewhat greater variability emerged in the smaller classes. Together, these findings suggest that attrition was substantial, particularly in symptomatic classes, but was not strongly associated with baseline symptom severity within classes. Importantly, despite a potential attrition-related bias toward apparent symptom improvement, the moderate-yet-diverging class in the 3-class solution continued to show modest increases in OCS over time while PTSS gradually declined. This pattern suggests that the relative persistence of OCS following trauma exposure—and, more broadly, the stronger association between interpersonal trauma exposure and OCS emergence/chronicity relative to PTSS—is unlikely to be fully explained by selective dropout alone.

## Discussion

This study is the first to evaluate OCS trajectories or combined PTSS/OCS trajectories in a trauma-exposed sample, showing both overlap and differentiation from earlier research on separate PTSS and OCS trajectories. Namely, the diverging PTSS and OCS trajectory suggests unique pathways for these related yet distinct symptom clusters, even in the aftermath of trauma exposure. Given the stronger associations between interpersonal traumas (i.e., physical or sexual abuse) and OCS emergence and chronicity compared to non-interpersonal traumas (e.g., natural disasters),^3,5,6^ replication in a sample of interpersonal trauma-exposed individuals and with larger sample sizes is needed.

### Limitations

Our results should be interpreted in light of three key limitations. First, the modest sample size limited statistical power and may have reduced our ability to detect finer-grained heterogeneity in trajectories within the intermediate severity class. This possibility is consistent with the 4-class solution, which suggested potential subdivision of the intermediate group but showed reduced stability across bootstrap analyses. Second, attrition across waves was substantial, particularly within the more symptomatic classes, raising the possibility that later-wave trajectories may underestimate symptom persistence over time. Although baseline PTSS and OCS severity were generally comparable between retained and dropout participants within classes, selective attrition related to unmeasured factors cannot be ruled out. Notably, despite this potential bias toward apparent symptom improvement, the intermediate class in the 3-class solution continued to show modest increases in OCS over time, suggesting that the relative persistence of OCS following trauma exposure is unlikely to be fully explained by selective dropout alone. Lastly, the lack of symptom severity data prior to Hurricane Helene significantly limits our ability to interpret the causal effects of trauma exposure on PTSS and OCS symptom trajectories. A meaningful proportion of cases in the symptomatic classes likely had clinically significant PTSS or OCS before Hurricane Helene. Their symptoms may have been unaffected, worsened, or even improved by trauma exposure; though the latter is unlikely, particularly for PTSS. Future research on posttraumatic symptom trajectories will need to evaluate and account for symptom severity prior to the index event to strengthen causal interpretations of the symptom trajectory analyses.

## Data Availability

All data produced in the present study are available upon reasonable request to the authors

## Acknowledgements

With profound gratitude to the survivors of Hurricane Helene who volunteered their time, energy, and resources to this study, in the hope of helping others.

